# Trends in Perioperative Catheter Utilization by Surgical Procedure Type in the United States, 2010 – 2017: Evidence from the Nationwide Inpatient Sample

**DOI:** 10.1101/2024.03.22.24304038

**Authors:** Priscilla D. Anderton, Anthony C. Waddimba, Gerald O. Ogola, Michael A. Ramsay, Bryan Kim, Gregory R. Thoreson

**Affiliations:** Texas A&M University, College of Medicine, College Station, Texas; Baylor University Medical Center, Department of Surgery, Division of Surgical Research, Dallas, Texas; Baylor Scott and White Research Institute, Dallas, Texas; Baylor University Medical Center, Department of Anesthesiology and Pain Management, Dallas, Texas; Baylor of University Medical Center, Department of Surgery, Division of Urology, Dallas, Texas; Department of Clinical Research, Urology Clinics of North Texas, Dallas, Texas; Adjunct Assistant Professor, Department of Engineering, University of Texas, Dallas, TX

**Author notes:** **Correspondence**: Gregory R. Thoreson, MD, FACS, Department of Surgery, Division of Urology, Baylor University Medical Center, 3417 Gaston Ave, Suite 830 Dallas, TX 35246.

**Keywords:** Indwelling urethral catheter, perioperative urethral catheterization, National Inpatient Sample, clinical practice variation, Mann-Kendall trend test, Locally estimated scatterplot smoothing

## Abstract

**Background:** Indwelling urethral catheters are regularly used in the operative setting to monitor urinary output, decompress the bladder, or manage post-operative urinary retention. Indwelling urethral catheterization can be associated with significant complications, such as nosocomial infections and urethral trauma, hence the contemporary national and institutional efforts to reduce indwelling urethral catheter usage.

**Objectives:** The purpose of this study is to investigate national trends in perioperative catheter usage across surgical procedure categories.

**Methods:** This is a retrospective study of adults who received indwelling urethral catheters during elective surgical procedures performed in United States (U.S.) hospitals during 2010-2017. We utilized the National Inpatient Sample (NIS) procedure codes to identify surgical procedure types and insertions of indwelling urethral catheters. Yearly catheterization rates were utilized to assess significant differences in nationwide temporal trends.

**Results:** We sampled 81,128,725 perioperative catheterizations across twelve procedure categories. When specific categories were examined separately, the annual proportion of procedures utilizing catheterization *decreased* for eye (−0.05% [95% CI, -0.10%; -0.02%], p-value 0.013), musculoskeletal (−0.04% [95% CI, -0.09%; 0.003%] p-value 0.063), and urinary system surgeries (−0.54% [95% CI, -0.76%; -0.33%] p-value 0.001). The proportion that utilized urethral catheters *increased* annually for female reproductive (+0.05% [95% CI, 0.04%; 0.07%] p-value <0.001), male reproductive (+0.31% [95% CI, 0.18%; 0.44%] p-value 0.001), and respiratory system surgeries (0.08% [95% CI, 0.02%; 0.13%] p-value 0.015). When all operative procedures were examined together, the overall annual proportion involving urethral catheterization did not significantly vary during 2010-2017.

**Conclusion:** Despite wide dissemination of clinical guidelines and institutional efforts to reduce catheterizations, we observed no significant changes in the overall proportion of all surgical procedures (irrespective of procedure category) that utilized urethral catheters during our study period.

## Introduction

The United States (U.S.) uses 30 million urinary catheters annually, with roughly $500 million spent on treating hospital-acquired urinary tract infections – a common complication of indwelling urethral catheter usage.^1,2^ In 2009, the Centers for Disease Control (CDC) promulgated clinical guidelines for appropriate use of catheters in the perioperative setting in an attempt to reduce unwarranted catheterization, associated complications, and costs.^2,3^ Little is known regarding nationwide trends in perioperative catheter usage, or how the proportion of surgical operations involving urethral catheterization has evolved since guideline dissemination.^4,5^ The present study queried the Nationwide Inpatient Sample (NIS) database to identify national yearly trends and annual percentage changes in perioperative catheter usage across surgical procedure categories.

### Methods

We analyzed elective surgical procedures performed with indwelling urethral catheters at U.S. hospitals during 2010-2017 and recorded in the NIS of the Agency for Health Research and Quality’s Healthcare Cost and Utilization Project (**eMethods** in the Supplement). The NIS list of procedure codes was used to identify the specific type of surgical procedure and the insertion of an indwelling urethral catheter (**eTables** in the Supplement). Only patients ≥ 18 years old were sampled.

The primary outcome was the proportion of surgical procedures involving indwelling urethral catheters. This was derived by dividing the count of adults with both a primary code for a surgical procedure and a secondary code for indwelling catheterization by the overall count of adults with primary codes for surgical procedures in each calendar year. Preliminary assessments of variations in annual urethral catheter utilization were based on the Chi-square test of proportions. Significance was defined at P < .05. Yearly catheterization rates were then configured as time series data. The Mann-Kendall trend test was applied to assess statistically significant differences in nationwide temporal trends in the derived time series. Locally estimated scatterplot smoothing curves were fitted to assess time series relationships. Statistical analyses were conducted using SAS® software version 9.4.

## Results

Between 2010 and 2017, a total of 81,128,725 perioperative catheterizations were performed in the U.S. Among twelve procedure groups, six demonstrated a significant change in proportion of catheters utilized annually. Annual catheter usage decreased at a rate of −0.05% ([95% CI, - 0.10%; -0.02%], p-value 0.013) for eye surgeries, −0.04% ([95% CI, -0.09%; 0.003%] p-value 0.063) for musculoskeletal system procedures, and −0.54% ([95% CI, -0.76%; -0.33%] p-value 0.001) for urinary system procedures (see **Table 1**; **Figure 1A**).

**Table 1.** Trends in indwelling urethral catheterization, 2010-2017.

| **Table 1. Trends in indwelling urethral catheterization, 2010-2017** |  |  |  |
| --- | --- | --- | --- |
| **Operating procedure** | **Direction of trend in catheterization 2010-2017** | **Rate of change** | **P-value** |
| Cardiovascular | Increasing | - | 0.966 |
| Ear, Nose, Mouth and Throat | Decreasing | - | 0.945 |
| Endocrine system | Increasing | - | 0.224 |
| *Eye* | *Decreasing* | -0.05% (-0.10%; -0.02%) | 0.013 |
| *Female reproductive* | *Increasing* | 0.05% (0.04%; 0.07%) | <0.001 |
| Gastrointestinal System | Increasing | - | 0.704 |
| Lymphatic and hemic system | Decreasing | - | 0.390 |
| *Male reproductive* | *Increasing* | 0.31% (0.18%; 0.44%) | 0.001 |
| Musculoskeletal system | Decreasing | -0.04% (-0.09%; 0.003%) | 0.063 |
| Nervous system | Decreasing | - | 0.097 |
| *Respiratory system* | *Increasing* | 0.08% (0.02%; 0.13%) | 0.015 |
| *Urinary system* | *Decreasing* | -0.54% (-0.76%; -0.33%) | 0.001 |
| All procedures | Decreasing | -0.0021 (-0.032, 0.028) | 0.873 |

Trends and rates in catheterization. Significance is determined from 95% confidence interval (CI).

**Figure 1.**
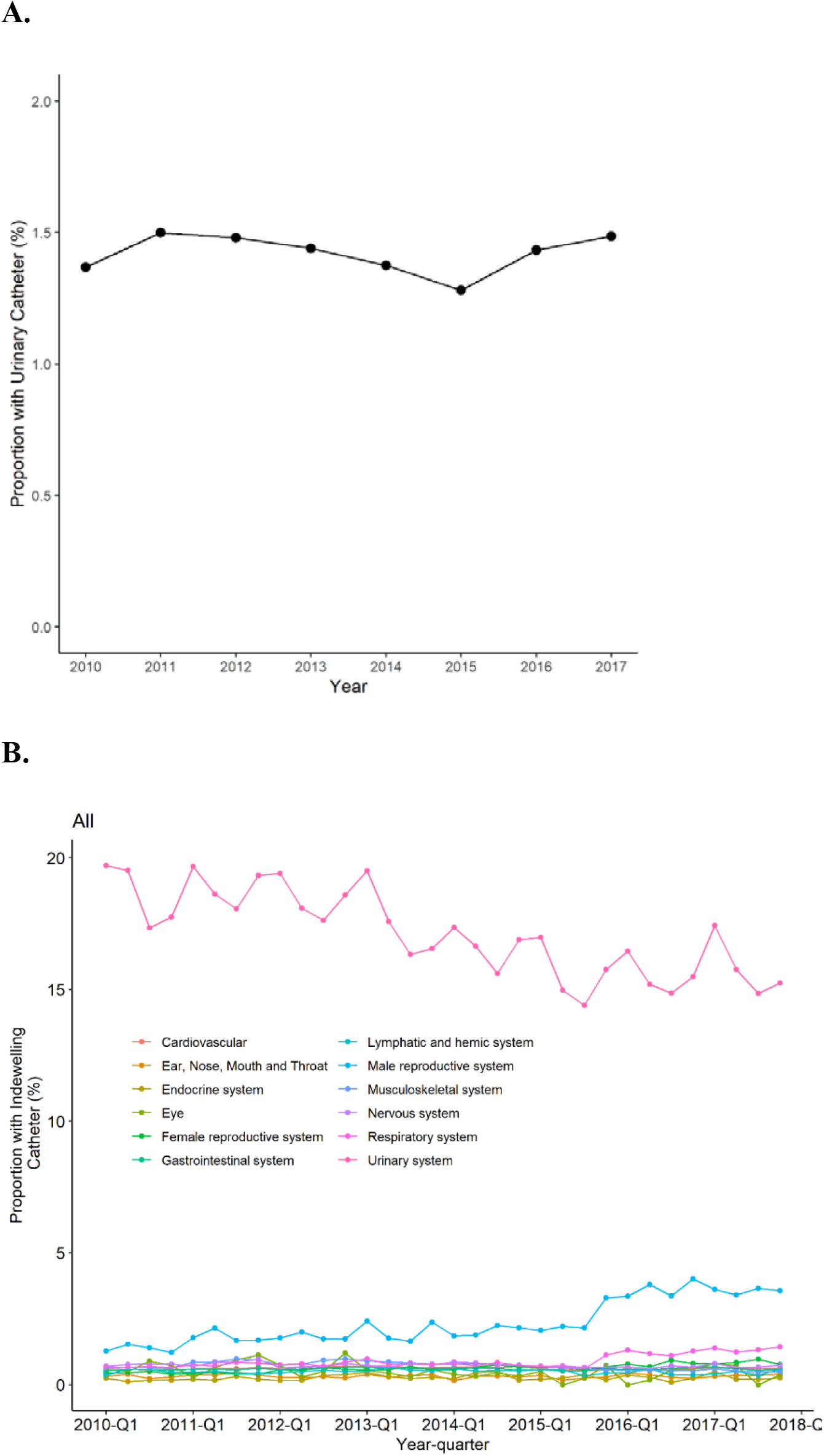
Proportion of Indwelling Catheters Utilized Between 2010 – 2017. **A**. **A:** Proportion of indwelling catheters utilized overall **B:** Proportion of indwelling catheters utilized within individual procedure groups

In contrast, catheter usage increased annually at a rate of +0.05% ([95% CI, 0.04%; 0.07%] p-value <0.001) for female reproductive procedures, +0.31% ([95% CI, 0.18%; 0.44%] p-value 0.001) for male reproductive procedures, and +0.08% ([95% CI, 0.02%; 0.13%] p-value 0.015) for respiratory system procedures (**Table 1**; **Figure 1A**). When all procedure groups were combined, there was no significant difference in the annual proportion of catheters used (annual trend, -0.0021% [95% CI, -0.032%; 0.028%]; p-value 0.873) (**Table 1**; **Figure 1B**).

## Discussion

Between 2010 and 2017, three procedure categories demonstrated a significant decrease in the annual proportion of catheters used in the perioperative setting, while three other procedure categories demonstrated a significant increase. However, when all operative procedures are considered together, the overall proportion of catheterizations did not significantly change during this period, despite the dissemination of CDC guidelines and reports of successful initiatives to reduce urinary catheterizations within select institutions.^5^ There are a few plausible explanations for this observation. This could be due to institution-level variation in guideline implementation or adherence.^4,5^ Additionally, this could be explained by the lack of a standardized alternative method for the accurate measurement of intraoperative urinary output – a common indication for catheterization.^6,7^ Thus, additional research studies are warranted to explore practical alternatives to indwelling urinary catheters that could be implemented in the ‘real world’ perioperative setting.

The present study has notable limitations. First, this is a retrospective study limited to information available within the NIS database. Secondly, biases resulting from periodic changes in the sampling strategy utilized during NIS data collection (**eMethods** in the Supplement) are possible.

## Supporting information

Supplemental Online Content

## Data Availability

All data produced in the present study are available upon reasonable request to the authors.

