## Supplemental Online Content for "Trends in Perioperative Catheter Utilization by Surgical Procedure Type in the United States, 2010 – 2017: Evidence from the Nationwide Inpatient Sample"

eMethods.

eTables. Diagnostic Codes Used to Define Operative Procedure Type and Urethral Catheterization Based on ICD-9 and ICD-10 Codes

eReferences.

This information was provided by the authors to give readers more information about their work.

**eMethods**

***1. Nationwide Inpatient Sample***

The Nationwide Inpatient Sample (NIS) database is a part of the Health Care Utilization Project (HCUP) and is maintained by the Agency for Healthcare Research and Quality (AHRQ). It provides a representative national sample of inpatients, which can be used to estimate nationwide hospitalization rates for different medical conditions.^1^ The NIS is sampled from the State Inpatient Databases (SID), which includes all inpatient data currently contributed to the HCUP.^2^ There are 47 states that participate in the US, plus the District of Columbia, which covers more than 97% of the US population. NIS data are available from 1988 through 2021, which allows analysis of trends over time. The NIS file structure consists of three discharge-level files (core, severity, and diagnosis and procedure groups) and one hospital-level file. The core file contains commonly used data elements, such as demographics and ICD-10-CM codes. The severity file contains additional data elements that aid in identifying the severity of the condition for a specific discharge. The diagnosis and procedure groups file contain additional information on ICD diagnoses and procedure codes provided by the AHRQ. The hospital file contains information on hospital-level characteristics.^1^

The NIS database has undergone numerous changes over time, including the sampling and weighting strategies used. In 2012, the NIS was redesigned to improve national estimates. These changes included revisions to the sample design, how hospital institutions are defined, and other revisions intended to enhance confidentiality. Details of these revisions can be found on the NIS [website](https://hcup-us.ahrq.gov/nisoverview.jsp). Additionally, until 2015, the NIS included a full calendar year of data with diagnosis and procedure codes reported using the ICD-9-CM codes. In October 2015, hospital administrative data began using the ICD-10-CM/PCS coding system.^1^ These changes must be considered when conducting longitudinal analyses.

***2. Identifying Operative Procedure and Indwelling Catheter Utilization***

We identified eligible cases by using ICD-9 procedure codes from 2010 up to the 3^rd^ quarter of 2015, and ICD-10 procedure codes from the 4^th^ quarter of 2015 up to 2017. The type of operating procedures was identified from primary ICD-9 or ICD-10 procedure type.  The ICD-9 and ICD-10 codes used to identify operative and surgical procedures are listed in **eTables** **A1** and **A2**, respectively. Instances of indwelling catheter utilization were identified using the secondary ICD-9 and ICD-10 procedure codes specified in Table B.^3^ We generated weighted hospitalization-level counts of patients with each code to derive estimates representative of the entire hospitalized population nationwide.

**eTables. Diagnostic Codes Used to Define Operative Procedure Type and Urethral Catheterization**

| **Table A1. ICD-9 surgical/operative procedures** | |
| --- | --- |
| **ICD-9 code** | **Procedure description** |
| 01.01  05.9 | 1. Operations on the nervous system |
| 06.01  07.99 | 2. Operations on the endocrine system |
| 08.01  16.99 | 3. Operations on the eye |
| 18.01  20.99 | 4. Operations on the ear |
| [21.00](https://www.findacode.com/code.php?set=ICD9V3&c=21.00)  [29.99](https://www.findacode.com/code.php?set=ICD9V3&c=29.99) | 5. Operations on the nose, mouth, and pharynx |
| [30.01](https://www.findacode.com/code.php?set=ICD9V3&c=30.01)  [34.99](https://www.findacode.com/code.php?set=ICD9V3&c=34.99) | 6. Operations on the respiratory system |
| [35.00](https://www.findacode.com/code.php?set=ICD9V3&c=35.00) - [39.99](https://www.findacode.com/code.php?set=ICD9V3&c=39.99) | 7. Operations on the cardiovascular system |
| [40.0](https://www.findacode.com/code.php?set=ICD9V3&c=40.0)  [41.99](https://www.findacode.com/code.php?set=ICD9V3&c=41.99) | 8. Operations on the hemic and lymphatic system |
| [42.01](https://www.findacode.com/code.php?set=ICD9V3&c=42.01)  [54.99](https://www.findacode.com/code.php?set=ICD9V3&c=54.99) | 9. Operations on the digestive system |
| [55.01](https://www.findacode.com/code.php?set=ICD9V3&c=55.01)  [59.99](https://www.findacode.com/code.php?set=ICD9V3&c=59.99) | 10. Operations on the urinary system |
| [60.0](https://www.findacode.com/code.php?set=ICD9V3&c=60.0)  [64.99](https://www.findacode.com/code.php?set=ICD9V3&c=64.99) | 11. Operations on the male genital organs |
| [65.01](https://www.findacode.com/code.php?set=ICD9V3&c=65.01)  [71.9](https://www.findacode.com/code.php?set=ICD9V3&c=71.9) | 12. Operations on the female genital organs |
| [76.01](https://www.findacode.com/code.php?set=ICD9V3&c=76.01)  [84.99](https://www.findacode.com/code.php?set=ICD9V3&c=84.99) | 14. Operations on the musculoskeletal system |
| [85.0](https://www.findacode.com/code.php?set=ICD9V3&c=85.0)  [86.99](https://www.findacode.com/code.php?set=ICD9V3&c=86.99) | 15. Operations on the integumentary system |

| **Table A2. ICD-10 surgical/operative procedures** | |
| --- | --- |
| **ICD-10 code** | **Procedure description** |
| [0016070](https://www.findacode.com/code.php?set=ICD10PCS&c=0016070)  [00XS4ZS](https://www.findacode.com/code.php?set=ICD10PCS&c=00XS4ZS) | [Body System 0 - Central Nervous System and Cranial Nerves](https://www.findacode.com/code-set.php?set=ICD10PCS&i=9798) |
| [012YX0Z](https://www.findacode.com/code.php?set=ICD10PCS&c=012YX0Z)  [01XH4ZH](https://www.findacode.com/code.php?set=ICD10PCS&c=01XH4ZH) | [Body System 1 - Peripheral Nervous System](https://www.findacode.com/code-set.php?set=ICD10PCS&i=10080) |
| [0210083](https://www.findacode.com/code.php?set=ICD10PCS&c=0210083)  [02YA0Z2](https://www.findacode.com/code.php?set=ICD10PCS&c=02YA0Z2) | [Body System 2 - Heart and Great Vessels](https://www.findacode.com/code-set.php?set=ICD10PCS&i=10324) |
| [0312090](https://www.findacode.com/code.php?set=ICD10PCS&c=0312090)  [03WYXMZ](https://www.findacode.com/code.php?set=ICD10PCS&c=03WYXMZ) | [Body System 3 - Upper Arteries](https://www.findacode.com/code-set.php?set=ICD10PCS&i=10604) |
| [0410090](https://www.findacode.com/code.php?set=ICD10PCS&c=0410090)  [04WYXKZ](https://www.findacode.com/code.php?set=ICD10PCS&c=04WYXKZ) | [Body System 4 - Lower Arteries](https://www.findacode.com/code-set.php?set=ICD10PCS&i=11072) |
| [051007Y](https://www.findacode.com/code.php?set=ICD10PCS&c=051007Y)  [05WYXKZ](https://www.findacode.com/code.php?set=ICD10PCS&c=05WYXKZ) | [Body System 5 - Upper Veins](https://www.findacode.com/code-set.php?set=ICD10PCS&i=11627) |
| [0610075](https://www.findacode.com/code.php?set=ICD10PCS&c=0610075)  [06WYXKZ](https://www.findacode.com/code.php?set=ICD10PCS&c=06WYXKZ) | [Body System 6 - Lower Veins](https://www.findacode.com/code-set.php?set=ICD10PCS&i=12019) |
| [072KX0Z](https://www.findacode.com/code.php?set=ICD10PCS&c=072KX0Z)  [07YP0Z2](https://www.findacode.com/code.php?set=ICD10PCS&c=07YP0Z2) | [Body System 7 - Lymphatic and Hemic Systems](https://www.findacode.com/code-set.php?set=ICD10PCS&i=12418) |
| [080N07Z](https://www.findacode.com/code.php?set=ICD10PCS&c=080N07Z)  [08XM3ZZ](https://www.findacode.com/code.php?set=ICD10PCS&c=08XM3ZZ) | [Body System 8 - Eye](https://www.findacode.com/code-set.php?set=ICD10PCS&i=12654) |
| [090007Z](https://www.findacode.com/code.php?set=ICD10PCS&c=090007Z)  [09WYX0Z](https://www.findacode.com/code.php?set=ICD10PCS&c=09WYX0Z) | [Body System 9 - Ear, Nose, Sinus](https://www.findacode.com/code-set.php?set=ICD10PCS&i=12979) |
| [0B110D6](https://www.findacode.com/code.php?set=ICD10PCS&c=0B110D6)  [0BYM0Z2](https://www.findacode.com/code.php?set=ICD10PCS&c=0BYM0Z2) | [Body System B - Respiratory System](https://www.findacode.com/code-set.php?set=ICD10PCS&i=13292) |
| [0C00X7Z](https://www.findacode.com/code.php?set=ICD10PCS&c=0C00X7Z)  [0CX7XZZ](https://www.findacode.com/code.php?set=ICD10PCS&c=0CX7XZZ) | [Body System C - Mouth and Throat](https://www.findacode.com/code-set.php?set=ICD10PCS&i=13594) |
| [0D11074](https://www.findacode.com/code.php?set=ICD10PCS&c=0D11074)  [0DYE0Z2](https://www.findacode.com/code.php?set=ICD10PCS&c=0DYE0Z2) | [Body System D - Gastrointestinal System](https://www.findacode.com/code-set.php?set=ICD10PCS&i=13855) |
| [0F140D3](https://www.findacode.com/code.php?set=ICD10PCS&c=0F140D3)  [0FYG0Z2](https://www.findacode.com/code.php?set=ICD10PCS&c=0FYG0Z2) | [Body System F - Hepatobiliary System and Pancreas](https://www.findacode.com/code-set.php?set=ICD10PCS&i=14357) |
| [0G20X0Z](https://www.findacode.com/code.php?set=ICD10PCS&c=0G20X0Z)  [0GWSX3Z](https://www.findacode.com/code.php?set=ICD10PCS&c=0GWSX3Z) | [Body System G - Endocrine System](https://www.findacode.com/code-set.php?set=ICD10PCS&i=14557) |
| [0H0T07Z](https://www.findacode.com/code.php?set=ICD10PCS&c=0H0T07Z)  [0HXNXZZ](https://www.findacode.com/code.php?set=ICD10PCS&c=0HXNXZZ) | [Body System H - Skin and Breast](https://www.findacode.com/code-set.php?set=ICD10PCS&i=14727) |
| [0J010ZZ](https://www.findacode.com/code.php?set=ICD10PCS&c=0J010ZZ)  [0JXR3ZZ](https://www.findacode.com/code.php?set=ICD10PCS&c=0JXR3ZZ) | [Body System J - Subcutaneous Tissue and Fascia](https://www.findacode.com/code-set.php?set=ICD10PCS&i=14986) |
| [0K2XX0Z](https://www.findacode.com/code.php?set=ICD10PCS&c=0K2XX0Z)  [0KXW4ZZ](https://www.findacode.com/code.php?set=ICD10PCS&c=0KXW4ZZ) | [Body System K - Muscles](https://www.findacode.com/code-set.php?set=ICD10PCS&i=15271) |
| [0L2XX0Z](https://www.findacode.com/code.php?set=ICD10PCS&c=0L2XX0Z)  [0LXW4ZZ](https://www.findacode.com/code.php?set=ICD10PCS&c=0LXW4ZZ) | [Body System L - Tendons](https://www.findacode.com/code-set.php?set=ICD10PCS&i=15629) |
| [0M2XX0Z](https://www.findacode.com/code.php?set=ICD10PCS&c=0M2XX0Z)  [0MXW4ZZ](https://www.findacode.com/code.php?set=ICD10PCS&c=0MXW4ZZ) | [Body System M - Bursae and Ligaments](https://www.findacode.com/code-set.php?set=ICD10PCS&i=16011) |
| [0N20X0Z](https://www.findacode.com/code.php?set=ICD10PCS&c=0N20X0Z)  [0NWWXMZ](https://www.findacode.com/code.php?set=ICD10PCS&c=0NWWXMZ) | [Body System N - Head and Facial Bones](https://www.findacode.com/code-set.php?set=ICD10PCS&i=16393) |
| [0P2YX0Z](https://www.findacode.com/code.php?set=ICD10PCS&c=0P2YX0Z)  [0PWYXMZ](https://www.findacode.com/code.php?set=ICD10PCS&c=0PWYXMZ) | [Body System P - Upper Bones](https://www.findacode.com/code-set.php?set=ICD10PCS&i=16712) |
| [0Q2YX0Z](https://www.findacode.com/code.php?set=ICD10PCS&c=0Q2YX0Z)  [0QWYXMZ](https://www.findacode.com/code.php?set=ICD10PCS&c=0QWYXMZ) | [Body System Q - Lower Bones](https://www.findacode.com/code-set.php?set=ICD10PCS&i=17083) |
| [0R2YX0Z](https://www.findacode.com/code.php?set=ICD10PCS&c=0R2YX0Z)  [0RWXXKZ](https://www.findacode.com/code.php?set=ICD10PCS&c=0RWXXKZ) | [Body System R - Upper Joints](https://www.findacode.com/code-set.php?set=ICD10PCS&i=17428) |
| [0S2YX0Z](https://www.findacode.com/code.php?set=ICD10PCS&c=0S2YX0Z)  [0SWWXJZ](https://www.findacode.com/code.php?set=ICD10PCS&c=0SWWXJZ) | [Body System S - Lower Joints](https://www.findacode.com/code-set.php?set=ICD10PCS&i=17870) |
| [0T13073](https://www.findacode.com/code.php?set=ICD10PCS&c=0T13073)  [0TY10Z2](https://www.findacode.com/code.php?set=ICD10PCS&c=0TY10Z2) | [Body System T - Urinary System](https://www.findacode.com/code-set.php?set=ICD10PCS&i=18233) |
| [0U15075](https://www.findacode.com/code.php?set=ICD10PCS&c=0U15075)  [0UY90Z2](https://www.findacode.com/code.php?set=ICD10PCS&c=0UY90Z2) | [Body System U - Female Reproductive System](https://www.findacode.com/code-set.php?set=ICD10PCS&i=18465) |
| [0V1N07J](https://www.findacode.com/code.php?set=ICD10PCS&c=0V1N07J)  [0VYS0Z2](https://www.findacode.com/code.php?set=ICD10PCS&c=0VYS0Z2) | [Body System V - Male Reproductive System](https://www.findacode.com/code-set.php?set=ICD10PCS&i=18666) |
| [0W0007Z](https://www.findacode.com/code.php?set=ICD10PCS&c=0W0007Z)  [0WY20Z1](https://www.findacode.com/code.php?set=ICD10PCS&c=0WY20Z1) | [Body System W - Anatomical Regions, General](https://www.findacode.com/code-set.php?set=ICD10PCS&i=18905) |
| [0X0207Z](https://www.findacode.com/code.php?set=ICD10PCS&c=0X0207Z)  [0XYK0Z1](https://www.findacode.com/code.php?set=ICD10PCS&c=0XYK0Z1) | [Body System X - Anatomical Regions, Upper Extremities](https://www.findacode.com/code-set.php?set=ICD10PCS&i=19150) |
| [0Y0007Z](https://www.findacode.com/code.php?set=ICD10PCS&c=0Y0007Z)  [0YWBXYZ](https://www.findacode.com/code.php?set=ICD10PCS&c=0YWBXYZ) | [Body System Y - Anatomical Regions, Lower Extremities](https://www.findacode.com/code-set.php?set=ICD10PCS&i=19338) |

| **Table B. ICD-9 and ICD-10 indicators for indwelling catheter utilization** | |
| --- | --- |
| **ICD-9 code** | **Procedure description** |
| 57.94 | Insertion of indwelling urinary catheter for and |
| **ICD-10 code** | **Procedure description** |
| 0T9B00Z | Drainage of Bladder with Drainage Device, Open Approach, |
| 0T9B30Z | Drainage of Bladder with Drainage Device, Percutaneous Approach, |
| 0T9B40Z | Drainage of Bladder with Drainage Device, Percutaneous Endoscopic Approach, |
| 0T9B70Z | Drainage of Bladder with Drainage Device, Via Natural or Artificial Opening, and |
| 0T9B80Z | Drainage of Bladder with Drainage Device, Via Natural or Artificial Opening Endoscopic |
